# Scan-Rescan Reliability of TRUST ASL for Simultaneous Measurement of Cerebral Blood Flow and Blood–Brain Barrier Water Exchange: A pilot study in Healthy Controls and Patients with Epilepsy

**DOI:** 10.64898/2026.04.21.26350807

**Authors:** Ning Hua, Lena Václavů, Joseph Sisto, Chad Farris, Osamu Sakai, Janine Barrett, Maria Stefanidou, Abrar Al-Faraj, Matthew Belisle, Muhammad M Qureshi, Ali Guermazi, Lee Goldstein, Sara K. Inati, Meher Juttukonda, Bruce Rosen, William Theodore, Matthias JP van Osch, Myriam Abdennadher

## Abstract

**Objective:** Water imbalance across the blood–brain barrier (BBB) can raise neuronal excitability and promote seizures. Noninvasive imaging of water exchange probes BBB function. T2-relaxation under-spin-tagging (TRUST) arterial spin labeling (ASL) separates intravascular and extravascular labeled water. We tested TRUST-ASL feasibility and scan–rescan reliability.

**Methods:** Twelve participants underwent two 3T MRI sessions 2–4 weeks apart. Unlike conventional TRUST for venous oximetry, in the present study TRUST refers to the incorporation of T2-preperation module within a time-encoded pCASL sequence for arterial labeled water. TRUST-ASL included 9 post labeling delays and 2 labeling delay. CBF and water exchange time (Tex) were quantified using MATLAB. Maps were registered to T1 and normalized to MNI space via FreeSurfer. Scan–rescan reliability was measured in grey matter and regions of interests using intraclass correlation coefficients (ICC). Level of agreement was evaluated with Bland–Altman plots and within-subject coefficient of variation. Statistical analyses used R.

**Results:** Scan-rescan reliability was good for cortical CBF (ICC = 0.79, 95% CI [0.28 - 0.94]) and excellent for Tex (ICC = 0.97, 95%CI [0.90-0.99]). Regional analyses revealed lower reliability in the basal ganglia compared to other subregions. Bland-Altman analysis demonstrated strong agreement between sessions, with mean differences near zero for CBF (limits of agreement: -20 to 20 ml/100g/min), and Tex (mean differences 0.17s; limits of agreement: -0.44 to 0.79 s).

**Conclusion:** Our study showed that TRUST-ASL as a promising chemical-free biomarker for group-level longitudinal studies of BBB dysfunction in epilepsy, while advising caution for individual-level longitudinal monitoring.

**HIGHLIGHTS:**

- TRUST-ASL is distinct from conventional TRUST MRI, enabling regional imaging rather than a single global venous oxygenation measurement.
- TRUST-ASL simultaneously measures cerebral blood flow and blood–brain barrier water exchange in vivo.
- CBF has high scan–rescan reliability but notable variability at the individual level.
- Tex has high relative reliability (ICC) in group analysis but substantial within-subject variability (wCV), suggesting caution during the interpretation of longitudinal changes especially at the individual level.

## INTRODUCTION

Epilepsy affects over 50 million people worldwide, posing a significant burden through recurrent seizures, comorbidities, and social isolation (1). Emerging research aims to elucidate mechanisms contributing to seizure recurrence and medication resistance, among which the blood-brain barrier (BBB) dysfunction has attracted increasing attention (2–9). The BBB is a highly selective neurovascular interface composed primarily of endothelial cells interconnected by tight junctions, surrounded by pericytes and astrocyte end-feet (10,11). Together, these components maintain a mechanical barrier and regulate selective transport between the bloodstream and brain parenchyma.

Water homeostasis across the BBB is primarily regulated by aquaporins expressed at astrocyte end-feet and is essential for normal neuronal function (12). Disruption of water balance can increase neuronal excitability and facilitate seizure generation (13–15). Previous epilepsy research has employed dynamic contrast-enhanced MRI (DCE-MRI) and other water tracers to quantify BBB permeability (16–20). However, subtle BBB leakages, typical in epilepsy due to focal and less overt barrier disruptions, pose challenges to the sensitivity of these methods. Moreover, evidence of intracranial gadolinium accumulation after contrast-enhanced MRI prompted safety reviews by the FDA and led the European Medicines Agency to restrict the use of certain linear agents to reduce potential risks to the brain, and suggests caution in the use of gadolinium-based contrast agents, especially for longitudinal monitoring (21–24).

Arterial spin labeling (ASL) MRI is a non-contrast technique using endogenous water protons as an almost freely diffusible “label.” It is widely employed to measure cerebral blood flow (CBF) in various neurological and psychiatric disorders including stroke, dementia, and epilepsy (25). Beyond perfusion quantification, advanced ASL-based methods can assess BBB functioning by tracking water exchange from the intravascular into extravascular compartments (14,26–28).

T2 relaxation under spin-tagging (TRUST) ASL MRI is an ASL-based approach that labels <u>arterial</u> water protons in the neck region, tracking their signal after arrival in brain tissue at multiple post-labeling delays (29,30). Unlike conventional TRUST MRI, which provides a single global measurement of venous blood oxygenation,(31) TRUST ASL combines Hadamard-encoded pCASL with a multi-TE TRUST module to generate voxel-wise maps of BBB water exchange, enabling regional quantification throughout the brain. Our implementation follows Petitclerc et al.(29) with a Hadamard-4 encoding scheme (instead of a Hadamard-8). Using a two-compartment model, this method allows measurement of CBF, arterial transit time (ATT), and BBB water exchange time (Tex) (29,32). While single-PLD ASL has been used to study CBF in epilepsy and multi-PLD ASL has been applied in aging and cerebrovascular research (33–37), and more recently by our team in epilepsy (32), few studies have evaluated the scan–rescan reliability of ASL metrics (38–42). In addition, chronic, interictal BBB dysfunction–distinct from ictal BBB opening–may relate to seizure severity, treatment response, and disease state (2,43–45).

Building on this, our study aim was to investigate the feasibility and inter-visit scan-rescan reliability of TRUST-ASL MRI in subjects with epilepsy and healthy controls. We hypothesized that this approach would allow reliable monitoring of physiologic CBF and BBB water exchange at least on a group level, but maybe even on the single subject level.

Given that TRUST-ASL MRI assesses BBB function noninvasively–without injection of agents–it is a promising tool for longitudinal monitoring of dynamic BBB changes not only in epilepsy, but also in other neurological conditions such as Alzheimer’s disease, traumatic brain injury, and stroke. In this pilot study, we evaluated the feasibility and reliability of CBF and BBB water exchange measurements using TRUST MRI in individuals with epilepsy and healthy controls.

## METHODS

### Study Population

The study was approved by the institutional review board, and all participants provided written informed consent prior to enrollment. Participants were recruited between September 2022 and January 2024. We focused this analysis on participants who successfully completed two MRI sessions, separated by 2 to 4 weeks, enabling assessment of scan-rescan reliability. Epilepsy participants were identified by screening clinical records at our epilepsy center and met inclusion criteria: age ≥18 years and confirmed focal epilepsy. Exclusion criteria were pregnancy or unknown pregnancy status, MRI contraindications, brain tumor or vascular malformation, vagus nerve stimulator, prior brain surgery, or inability/unwillingness to complete study questionnaires. Healthy controls were recruited via screening questionnaire and met inclusion criteria: age ≥18 years and no history of neurologic, psychiatric, or other medical conditions affecting the central nervous system. After consent, participants were scheduled for an initial MRI visit and were offered the opportunity to return for a second scan 2 to 4 weeks later. All participants completed MRI safety screening prior to imaging. Additionally, epilepsy participants completed a detailed questionnaire before each scan session, capturing information on their most recent seizure, semiology to inform on seizure type following the International League Against Epilepsy (ILAE) 2025 seizure classification (46), and any medication changes since their last epilepsy clinic visit. Data from twelve participants (5 healthy controls and 7 individuals with epilepsy) who completed two brain MRI sessions were included in this analysis.

### Brain MRI acquisition

Brain MRI was acquired on a 3T Philips MR 7700 imaging system with a 32-channel head coil. Total examination duration was ∼30 minutes. Sequences included structural MRI with T1-weighted Magnetization-Prepared Rapid Gradient-Echo (MPRAGE), fluid attenuation inversion recovery (FLAIR), and TRUST-ASL. MPRAGE’s key parameters were: flip angle = 8°, repetition time (TR) = 6.7ms, echo time (TE) = 3.0ms, field of view (FOV) = 250x250x170mm^3^, resolution = 1x1x1mm^3^. FLAIR’s key parameters were: inversion time = 1650ms, TR = 4800ms, effective TE = 340ms, resolution = 0.625x0.625x0.56mm^3^. For TRUST sequence, we used a pCASL sequence with a Hybrid Hadamard-4 labeling scheme and a 3D GRASE (gradient and spin-echo) readout. Arterial blood labeling was performed on internal carotid arteries at the neck after internal/external carotid bifurcation with precise manual placement of labeling slab by an expert/trained research MRI technician (level is also approximately at C2-C3 vertebrae). Key parameters were: labeling durations (LD) (650ms, 650ms, 950ms, 650ms, 950ms, 950ms, 2000ms, 2000ms, 2000ms), corresponding post-labelling delays (PLD) (200ms, 650ms, 850ms, 900ms, 1300ms, 1550ms, 1800ms, 2250ms 2500ms), TR = 6000ms, TE = 12ms, field of view = 240x240x108 mm^3^, resolution = 3.75x3.75x6mm^3^, and a separate equilibrium magnetization (M0) image (TR = 2000ms, same readout) (Figure 1).

**Figure 1.**
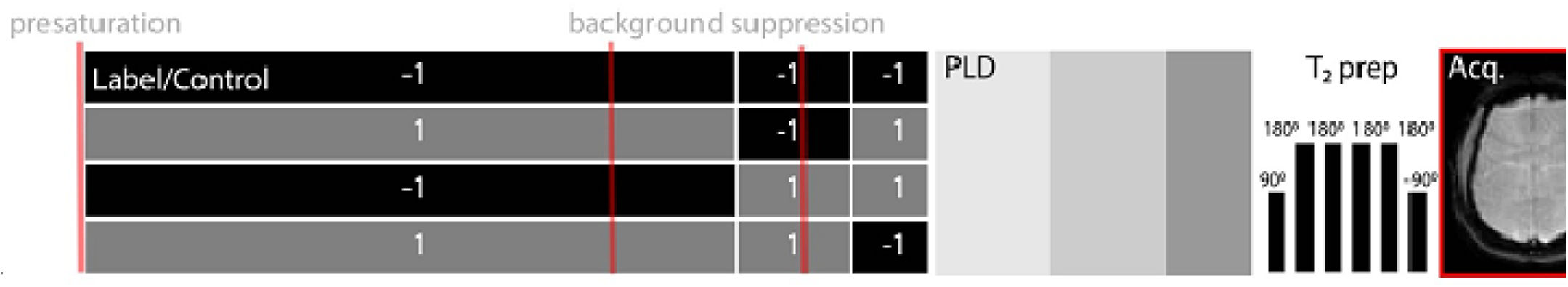
TRUST-ASL sequence diagram. Arterial blood labeling is performed on carotid arteries at the neck after internal/external carotid bifurcation (∼vertebrae C2-C3). Labeling/control was performed in a time-encoded fashion using a Hadamard-4 matrix to encode three labeling durations (LDs) per matrix. The sequence was repeated three times, each with a different post-labeling delay (PLD), resulting in 9 combinations of LD and PLD in total. In addition, a T2 preparation module (T2prep) applied before the 3D GRASE acquisition (Acq.) encodes four effective echo times per PLD. Pre-saturation and background suppression pulses are included to improve signal to noise ratio (SNR) and to reduce motion artifacts.

All data were checked for data quality and motion artifact. Structural images (T1-weighted and FLAIR) in healthy participants were visually reviewed by two neuroradiologists (OS, CF) to ensure that there was no incidental concerning lesion or destructive cortical lesion.

### Quantification of CBF and water exchange across the BBB (Tex)

ASL signal from 9 post-labeling delays and labeling duration pairs were extracted using Hadamard decoding. Quantification was performed in two steps. In the first step, CBF and arterial transit time (ATT) were estimated with in-house developed MATLAB software using least-squares fitting (MathWorks, MA) of the model described by Buxton et al (47). In the second step, CBF and ATT from the previous step were fixed and water exchange time (Tex) was quantified using a two-compartment approach to account for finite water exchange (29,48).

CBF, ATT, and Tex images were registered to participants’ structural T1-weighted image. FreeSurfer was used for registration and segmentation following the Desikan-Killiany Atlas (49). Regions of interest were grouped into temporal, frontal, basal ganglia, occipitoparietal, and all cortical grey matter after calculating weighted averages using R.

### Statistical analysis

We evaluated TRUST-ASL feasibility via scan-rescan comparisons of CBF, ATT, and Tex from two separate sessions. Reliability was assessed in cortical grey matter and four regions of interest (ROIs): basal ganglia (BG), temporal (including amygdala and hippocampus), frontal, and occipitoparietal cortices. For each ROI, we computed average values (per volume) of CBF, ATT, and Tex, and used complementary reproducibility measures.

We employed two-way mixed-effects intraclass correlations (ICCs) and chose to report both ICCsingle [3,1] for single-measure reliability and ICCaverage [3,k] for the mean of k=2 measurements because they quantify reliability under different measurement scenarios. ICCsingle (3,1) reflects the reliability of a single measurement, representing the reproducibility of TRUST scan if only one scan is acquired (useful for typical clinical indications). In contrast, ICCaverage (3,k) represents the reliability of the average of k repeated measurements (k = 2 in our study). Averaging multiple measurements typically reduces measurement error and results in higher reliability estimates (useful for planning longitudinal study designs or protocols relying on repeated assessments). ICC interpretation followed Koo & Li (50).

Bland-Altman analyses were conducted to evaluate agreement and systematic differences between scan sessions for each TRUST metric, providing insight into the measurement variability across repeated acquisitions (51). Within-subject coefficient of variation (wCV) was used to estimate each measurement variability within subjects. All analyses were performed in R.

These combined metrics provide both method reliability (ICC), precision and absolute agreement (wCV and Bland-Altman) for TRUST-derived CBF, ATT, and Tex.

## RESULTS

### Subject Characteristics

Twelve subjects completed two brain MRI sessions (n = 12; mean age = 42 ± 7 years old; range = 33-59; 7 women), including 5 healthy controls (43 ± 6 years old; 3 women) and 7 individuals with epilepsy (41± 9 years old; 4 women). Brain MRIs were completed within 4 weeks (mean interval = 23 ± 3 days; range [19-28]). Figure 2 shows T2 map and perfusion images obtained in a representative healthy subject. One epileptic participant had a focal seizure with preserved consciousness between the brain MRI sessions (at their usual seizure frequency) (Subject 2 in Supplemental Table 1).

**Figure 2.**
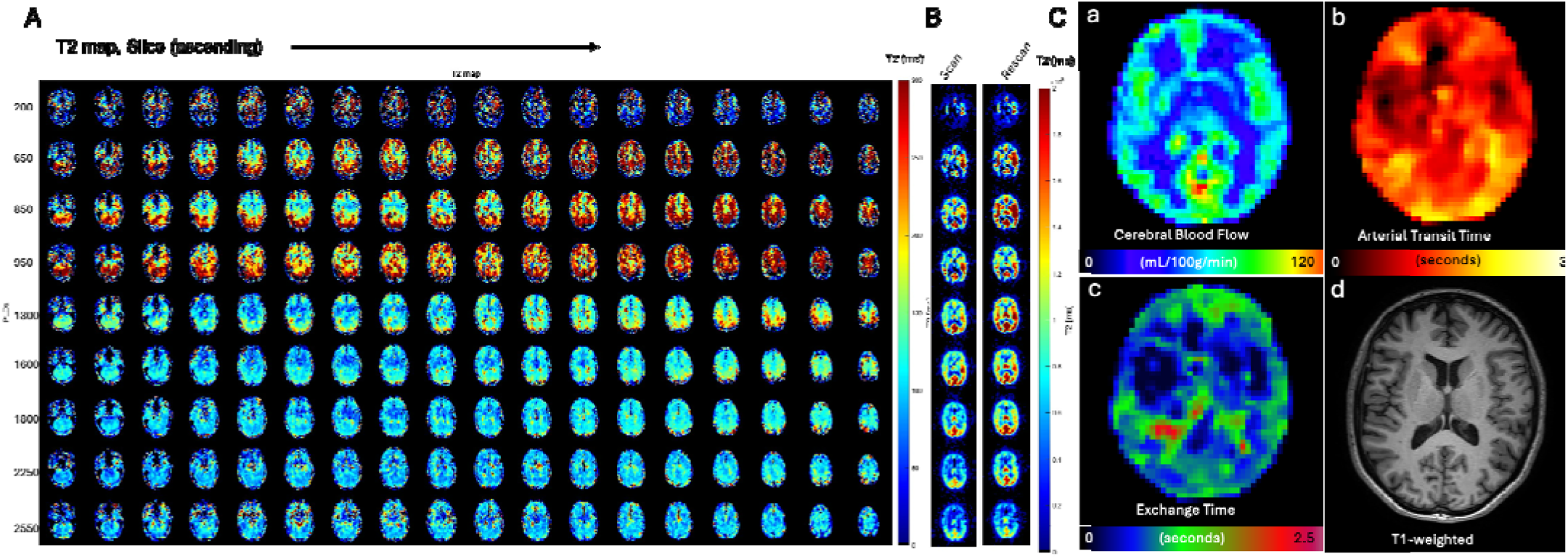
Representative images in a healthy participant. Figure shows T2 map of ASL-label with variable PLDs (A) and a selected slice scan and rescan T2 at 9 PLDs (B) (note that slices may not correspond perfectly in scan and rescan because images are not reregistered). (C) 3D images in the same healthy participant show measurements obtained from TRUST-ASL sequence. Using 2-compartment model outlined by Gregori et al (48), we quantified cerebral blood flow (CBF, b), arterial transit time (ATT, b), and water exchange time (Tex,c). T1-weighted image of the corresponding slice is shown (e).

### TRUST reliability using Intraclass correlation coefficient (ICC)

To evaluate the reliability of TRUST measurements we report ICCsingle [3,1] for single-measure reliability and ICCaverage [3,k] for the mean of k = 2 measurements. Regional scan-rescan reliability was good to excellent across all three measurements. Overall, Tex measurements exhibited superior scan-rescan reliability compared to CBF across all ROIs (Table 1).

**Table 1.** TRUST-ASL scan-rescan reliability. For each measure we summarize CBF, ATT, and Tex values in our study population. Then, we compared scan-to-scan reliability (ICC). We assessed reliability in all gray matter, basal ganglia, and cortical subregions: temporal, frontal, and parieto-occipital.

| Measure Location | ATT mean (SD) seconds |  | ATT reliability |  | CBF mean (SD) ml/100g/min |  | CBF reliability |  | Tex mean (SD) seconds |  | Tex reliability |  |
| --- | --- | --- | --- | --- | --- | --- | --- | --- | --- | --- | --- | --- |
|  | Visit 1 Mean(SD) | Visit 2 Mean(SD) | ICCaverage (95% CI) | ICCSingle (95% CI) | Visit 1 Mean(SD) | Visit 2 Mean(SD) | ICCaverage (95% CI) | ICCSingle (95% CI) | Visit 1 Mean(SD) | Visit 2 Mean(SD) | ICCaverage (95% CI) | ICCSingle (95% CI) |
| Temporal Cortex | 1.242 (0.25) | 1.183 (0.206) | **0.966** (0.881 - 0.99) | **0.934** (0.788 - 0.981) | 60.632 (16.159) | 60.787 (11.365) | 0.793 (0.282 - 0.94) | 0.657 (0.164 - 0.888) | 1.242 (0.250) | 1.183 (0.206) | 0.966 (0.881-0.990) | 0.934 (0.788-0.981) |
| Basal Ganglia | 1.012 (0.128) | 0.973 (0.12) | 0.876 (0.57 - 0.964) | 0.78 (0.398 - 0.931) | 55.42 (13.394) | 55.784 (9.279) | 0.712 (0 - 0.917) | 0.553 (0 - 0.847) | 1.012 (0.128) | 0.9730 (0.120) | 0.876 (0.570-0.964) | 0.790 (0.398 -0.931) |
| Frontal Cortex | 1.232 (0.174) | 1.171 (0.172) | **0.942** (0.797 - 0.983) | 0.889 (0.662 - 0.967) | 60.853 (16.21) | 61.782 (11.34) | 0.79 (0.269 - 0.939) | 0.652 (0.156 - 0.886) | 1.232 (0.174) | 1.171 (0.172) | 0.942 (0.797-0.983) | 0.889 (0.662-0.967) |
| **Occipitoparietal Cortex** | 1.469<br>(0.267) | 1.419<br>(0.25) | 0.972<br>(0.902-0.992) | 0.945<br>(0.821-0.984) | 68.187<br>(19.156) | 67.994<br>(13.422) | 0.795<br>(0.287-0.941) | 0.659<br>(0.168-0.888) | 1.469<br>(0.267) | 1.419<br>(0.250) | 0.972<br>(0.902-0.992) | 0.945<br>(0.821-0.984) |
| **All Grey Matter** | 1.311<br>(0.223) | 1.254<br>(0.205) | 0.97<br>(0.894-0.991) | 0.941<br>(0.809-0.983) | 63.816<br>(17.066) | 64.087<br>(11.934) | 0.791<br>(0.275-0.94) | 0.655<br>(0.159-0.887) | 1.311<br>(0.227) | 1.254<br>(0.205) | 0.970<br>(0.895-0.991) | 0.941<br>(0.809-0.983) |

### TRUST precision using Bland-Altman

To evaluate agreement and potential systematic bias between two scans, we performed Bland-Altman analysis. This method plots the differences against the averages of repeated measurements, with the mean difference indicating systematic bias (values near zero suggest minimal bias). The limits of agreement (LoA) represent the range containing 95% of differences, where narrower LoA indicate better agreement (52). The mean difference for ATT was 0.083 s, with LoA ranging from -0.10 to 0.21 s.

For CBF, the mean difference was -0.30 ml/100g/min, with upper LoA of -23.09 to 22.48ml/100g/min. Tex showed a mean difference of 0.17s, with LoA between -0.44 and 0.79 s.

Our study found minimal systematic bias for CBF, ATT, and Tex measurements obtained with the TRUST-ASL method across two scan sessions, indicating good agreement between sessions (Figure 3). The LoAs were narrow, and the small percentage differences between mean measurements (Table 2) suggest that variability between acquisition is unlikely to impact clinical interpretation.

**Figure 3.**
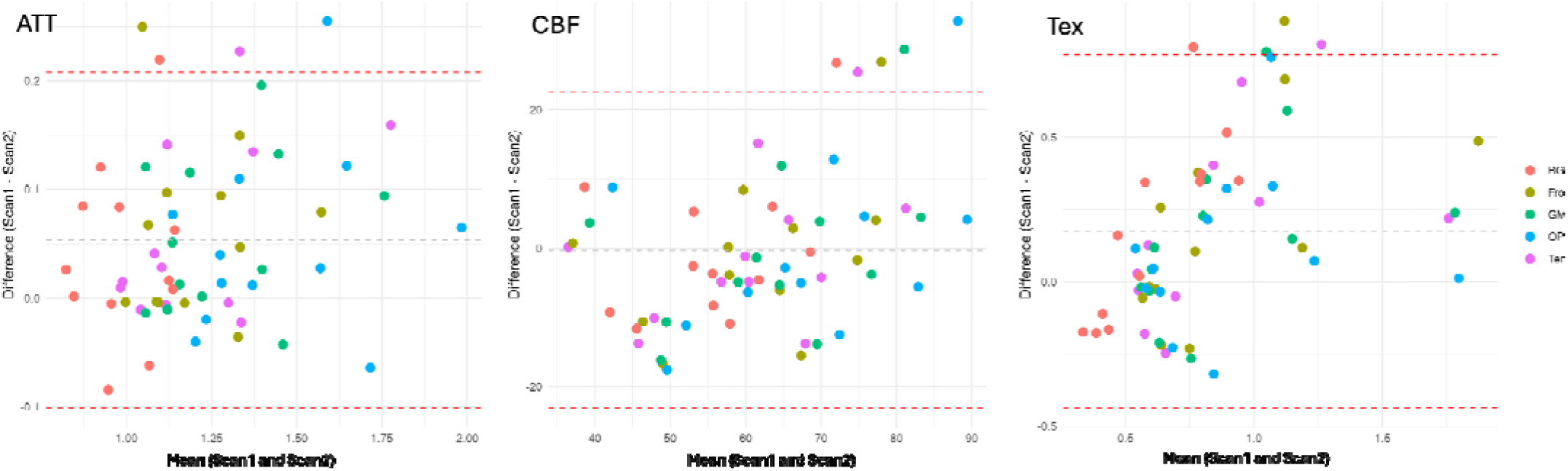
Bland-Altman plot of level of agreement in all regions of interest (ROI) of TRUST-ASL method quantification of CBF, ATT, and Tex. BG: basal ganglia (thalami, caudate, putamen, pallidum), GM: grey matter for all lateral cortex, OP: occipitoparietal cortex, Temporal: temporal cortex, hippocampi, and amygdala.

**Table 2.** Percent difference in mean measurements obtained at each TRUST MRI session

| Measure Region | ATT | CBF | Tex |
| --- | --- | --- | --- |
| **Temporal Cortex** | 2.4% | 0.10% | 2.40% |
| **Basal Ganglia** | 2% | 0.30% | 2% |
| **Frontal Cortex** | 2.50% | 0.80% | 2.40% |
| **Occipitoparietal Cortex** | 1.70% | 0.10% | 2.40% |
| **All Grey Matter** | 2.20% | 0.20% | 2.40% |

### Individual TRUST measurement consistency using within-subject coefficient of variation (wCV)

Following assessment of method consistency, we evaluated precision by calculating wCV. The wCV indicates the minimum detectable change, reflecting the sensitivity. We found higher wCV in Tex than in CBF and in CBF than in ATT (Table 3).

**Table 3.** Within subject coefficient of variation (wCV) in 12 participants.

| **Region of interest** | **wCV ATT (CI) %** | **wCV CBF (CI)%** | **wCV Tex (CI) %** |
| --- | --- | --- | --- |
| **Temporal Cortex** | 5.79 (4.15-9.56) | 12.90 (9.25-21.30) | 31.15 (22.34-51.42) |
| **Basal Ganglia** | 6.26 (4.49-10.33) | 13.27 (9.52-21.91) | 41.53 (29.79-68.57) |
| **Frontal Cortex** | 5.82 (4.17-9.60) | 12.92 (9.26-21.33) | 31.51 (22.60-52.01) |
| **Occipitoparietal cortex** | 4.70 (3.37-7.77) | 13.57 (9.73-22.40) | 23.18 (16.63-38.27) |
| **All Grey Matter** | 4.99 (3.58-8.23) | 12.96 (9.29-21.40) | 27.37 (19.63-45.18) |

## DISCUSSION

Our study showed feasibility of BBB water exchange measurement (Tex) with spatial mapping of BBB exchange in healthy and epileptic individuals. To our knowledge, this is among the first studies to investigate the feasibility of measuring BBB water exchange (Tex) and to assess scan-rescan reliability of CBF, ATT, and Tex measurements in epilepsy.

### Overall Method Reliability

Our primary finding demonstrates good to excellent CBF measurement reliability. While Tex demonstrates excellent relative reliability (high ICC), its comparatively high within-subject variability (wCV) limits sensitivity for detecting small longitudinal changes at the individual level (50–52). However, the relatively elevated wCV suggests imperfect precision potentially possibly due to sensitivity to physiological noise sources, including respiratory motion, cardiac pulsation, and cerebrospinal fluid dynamics (53,54).

We speculate that longer intervals between our scan sessions compared to other studies may have caused higher wCV and partly reflect true physiological variations. In fact, variability was smallest in intra-session wCV, reported by Mahroo et al (42) (6.6%) and Zhu et al (55) (8,10 and 15% for PLDs 2100, 1800, and 1500 ms respectively), compared to inter-session (7.9% by Mahroo et al), inter-visit with scans one week apart (8.4% by Mahroo et al), and inter-visit with scans 2 to 4 weeks apart in our study (24.9%). Further improving precision, either by accounting or controlling for key physiological factors (in future study design), is particularly critical for detecting small disease-related alterations in epilepsy, which may otherwise be obscured by physiological variation. In contrast, neurological disorders that induce more pronounced BBB disruption such as neurodegenerative diseases, traumatic brain injury, and stroke, yield changes sufficiently large to be reliably detected despite noise-related variability (56–59).

Our scan-rescan reliability analysis also showed that BBB water exchange measurements were more stable across repeated scans, compared to CBF in the group analysis (using ICC). But, measurements precision at the individual level quantified by wCV showed better precision in CBF than Tex using TRUST MRI, unlike previous work by Mahroo et al showing higher inter-visit wCV on CBF measurements compared to inter-visit wCV on Tex measurement (42), suggesting that BBB exchange measurements could represent a more robust biomarker for longitudinal studies than CBF or could be used in combination with CBF and other functional brain measurements.

### Region-wise findings

We found good-to-excellent scan-rescan reliability for ATT, CBF, and Tex across all ROIs, except BG-ROI where reliability was notably lower, particularly shown by the single measurement ICC. Additionally, CBF reliability in the frontal cortex was slightly lower but still considered good (ICC = 0.889). However, BG-ROI consistently exhibited the weakest reliability measures for all three parameters, with CBF showing the lowest ICC values. It is worthwhile noting measurements in this ROI showed shorter ATT and lower CBF on both scan sessions. So, this lower consistency in all three measurements in subcortical grey matter may be due to shorter ATTs in this region, or proximity to large vessels and ventricles. The association of shorter ATTs with better scan-rescan consistency (ICC) in BBB water exchange was also noted by recent work by Zhu et al (55). Furthermore, the basal ganglia receive blood from the proximal M1 segment of the middle cerebral artery near the circle of Willis; therefore, labeled protons in these nearby vessels may still bias our measurements despite corrections for macrovascular signal (60). Other ASL method-based studies also found limitations from measurements in basal ganglia / subcortical grey matter possibly due to basal ganglia’s relatively high iron content (affecting T1-blood), anatomical proximity to large vessels (macrovascular contamination), ventricles containing cerebrospinal fluid (partial volume effect), small size of the anatomical ROIs, transit time sensitivity, and the method’s low signal-to noise ratio (48,61–64).

We also noted longer exchange times in posterior regions. Although Tex and water exchange rate (Kw: defined as the capillary permeability surface area product of water, divided by distribution volume of water tracer in the capillary) are not directly comparable, previous studies have reported lower Kw in posterior regions—suggesting faster exchange—contrary to our findings (55).

### Impact on clinical care in epilepsy

The spatial extent of BBB alteration is an important consideration in epilepsy disease, especially for seizure-focus localization during the surgery evaluation. Studies have investigated BBB permeability during imaging work-up prior to epilepsy surgery by using DCE MRI (9), but this method requires the injection of exogenous substances potentially harmful to brain tissue and other organs (65–68). Several contrast-free techniques were developed using water as an endogenous label in ASL MRI to measure BBB water exchanges (14,27,29,42,48,69–76). Global BBB dysfunction in neurodegenerative conditions facilitate detection due to widespread involvement, whereas focal epilepsy involves localized BBB changes often limited to the seizure focus, or local regions. Combining a specific region of interest with longitudinal study designs may enhance the ability to monitor changes over time within focal areas (9,16,77–79).

### Limitations

Our pilot study demonstrates the feasibility of TRUST-ASL for measuring CBF and BBB water exchange, and provides preliminary scan-rescan reliability estimates, but the small sample and wide confidence intervals limits inference at the individual level; larger studies are needed to confirm clinical utility.

Our TRUST-ASL protocol used a T2-preparation module to separate intra- and extravascular ASL signal, but this reduces SNR at long effective echo times (29), and may be confounded by the oxygenation-dependent T2 changes across the vascular tree. Long ATTs can also bias quantification (80), and adding multiple T2 preparation times lengthens scans and increases motion sensitivity (81). Diffusion-weighted ASL is an alternative, though it relies on distinguishing compartments based on a single encoding velocity.

The cohort was small and evenly split between healthy controls and people with epilepsy, per expert panel recommendations. The sample size was further reduced because of the need to recruit participants for a second MRI within four weeks. Including epilepsy patients may have introduced signal variability from fluctuating neuronal activity and focal CBF changes; concurrent electroencephalogram (EEG) could be subclinical (no symptom) fluctuations, but MRI-compatible EEG for ASL may not be feasible because RF pulses increase tissue heating risk (82,83). We did not perform blood tests in the healthy participants; clinical records indicated no significant anemia (<10 g/dl) in epilepsy participants. Future studies should consider measuring hemoglobin/hematocrit for improved ASL quantification Finally, our 3–4-week scan-rescan interval exceeded the one-week intervals used in previous studies and could introduce within-subject variability from lifestyle factors (e.g. sleep, alcohol, smoking, caffeine) that affect CBF and BBB measures (84–88).

## CONCLUSION

Our pilot study demonstrates that TRUST MRI has good to excellent reliability (high ICC), although these results should be interpreted cautiously given the small sample size. TRUST-ASL is well suited for detecting group-level perfusion changes, but its current precision (wCV) may be inadequate for reliably detecting subtle within-subject alterations; the present data can inform sample size estimates for future longitudinal studies. Using machine-learning methods to reduce physiological noise and targetting region-specific BBB alterations could substantially improve biomarker precision and thereby enhance the utility of TRUST MRI for monitoring both diffuse and focal neurological disorders.

## Data Availability

Data is available upon reasonable request from the corresponding author.

## ACKNOWLEDGEMENTS

This study was funded by Boston University Clinical and Translational Science Institute (CTSI) Pilot Grant Program (1UL1TR001430).

## ETHICAL PUBLICATION STATEMENT

We confirm that we have read the Journal’s position on issues involved in ethical publication and affirm that this report is consistent with those guidelines.

## ABBREVIATIONS

ATT: arterial transit time
FUS: focal unconscious seizure
FCS: focal conscious seizure. LD, labeling delay
pCASL: pseudo-continuous arterial spin labeling
PLD: post-labeling delay
SD: standard deviation
Tex: exchange time.

