## Supplemental for "Scan-Rescan Reliability of TRUST ASL for Simultaneous Measurement of Cerebral Blood Flow and Blood–Brain Barrier Water Exchange: A pilot study in Healthy Controls and Patients with Epilepsy"

Supplemental table 1: Clinical and scan information of epilepsy participants

| **ID** | **Age range**  **(years)** | **Sex** | **Epilepsy**  **Duration**  **(years)** | **Type** | **Seizure Frequency** | **Interval from last seizure** | **Interval between visits**  **(days)** | **Seizure between visits** | **Medications** | **Seizure focus** |
| --- | --- | --- | --- | --- | --- | --- | --- | --- | --- | --- |
| 1 | 41-50 | M | 17 | FIC | every 2months | 4 days | 23 | no | CBZ, TPX, LMT | LT (left hippocampus and temporal tip confirmed on SEEG) |
| 2 | 31-40 | F | 16 | FPC*, FIC, FBTC | FM multiple per week, FUS and BTC 1 to 2 per month | FIC 17 days prior, FPC 1 Day prior | 28 | yes, FBTC 9 days prior, FPC 1 day | CNB, LCS, OXC | LF (inferior frontal gyrus confirmed on SEEG) |
| 3 | 31-40 | M | 25 | FPC, FIC, FBTC | none | last FPC >13 months, last FIC and FBTC>20years | 19 | no | LEV | LT |
| 4 | 31-40 | F | 7 | FIC | none | 7.5 years | 21 | no | OXC | LT |
| 5 | 51-60 | F | 11 | FIC | none | 9.5 years | 22 | no | LEV | BiT |
| 6 | 41-50 | F | 35 | FIC | none | 16 years | 21 | no | VPA | LT |
| 7 | 31-40 | M | 28 | FIC | every 4 months | 6 months | 21 | no | CNB, LEV, OXC | RT (broad lateral temporal cortex) |
| FBTC: Focal to bilateral tonic clonic seizure, FIC: Focal impaired consciousness seizure, FPC: Focal preserved consciousness seizure (FPC), *dystonic, LT: Left temporal, RT: Right temporal, BiT: Bilateral temporal, LF: Left frontal, CBZ: Carbamazepine, CNB: Cenobamate, LCS: Lacosamide, LEV: Levetiracetam, LMT: Lamotrigine, OXC: Oxcarbazepine, TPX: Topiramate, VPA: Valproic acid, SEEG: Stereo EEG (intracranial EEG recording). Seizure type was captured following the 2025 ILAE seizure classification (Beniczky et al, Epilepsia 2025). | | | | | | | | | | |

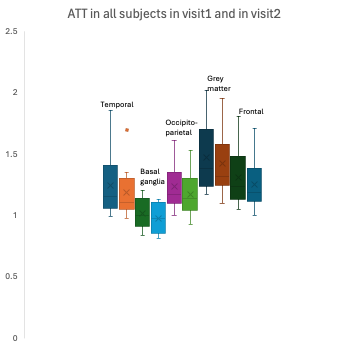

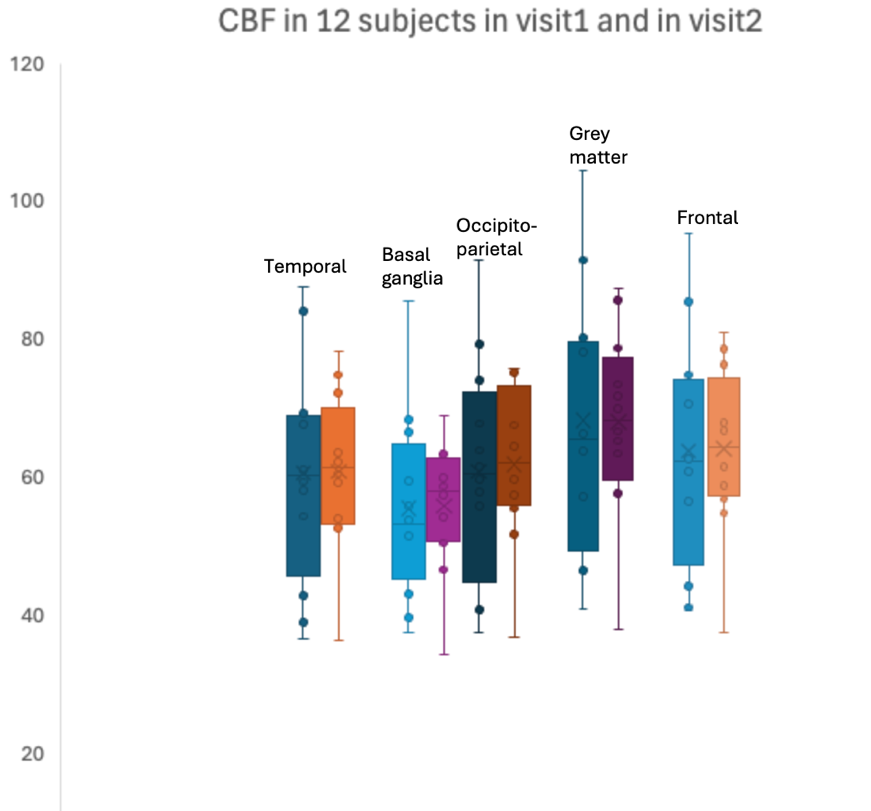

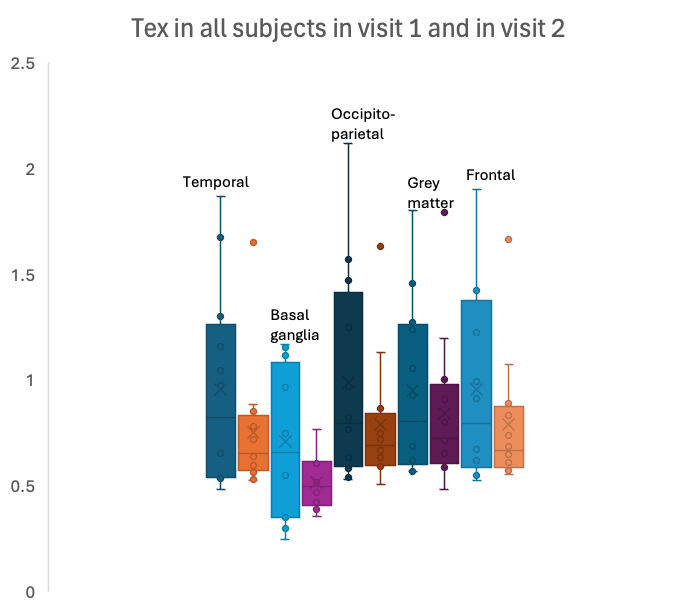
**Supplemental Figure 1.** TRUST measurements values at each region of interest (ROI)

**Supplemental Figure 2.** Individual Bland-Altman in each subject at each region of interest (ROI) for each TRUST measurement. Panel A shows Bland-Altman plot for cerebral blood flow (CBF) in each ROI. Panel B shows Bland-Altman plot for water exchange time (Tex blood-tissue) in each region of Interest. Panel C shows Bland-Altman plot for arterial transit time (ATT) in each ROI. LoA, lower level of agreement, UoA, upper level of agreement.

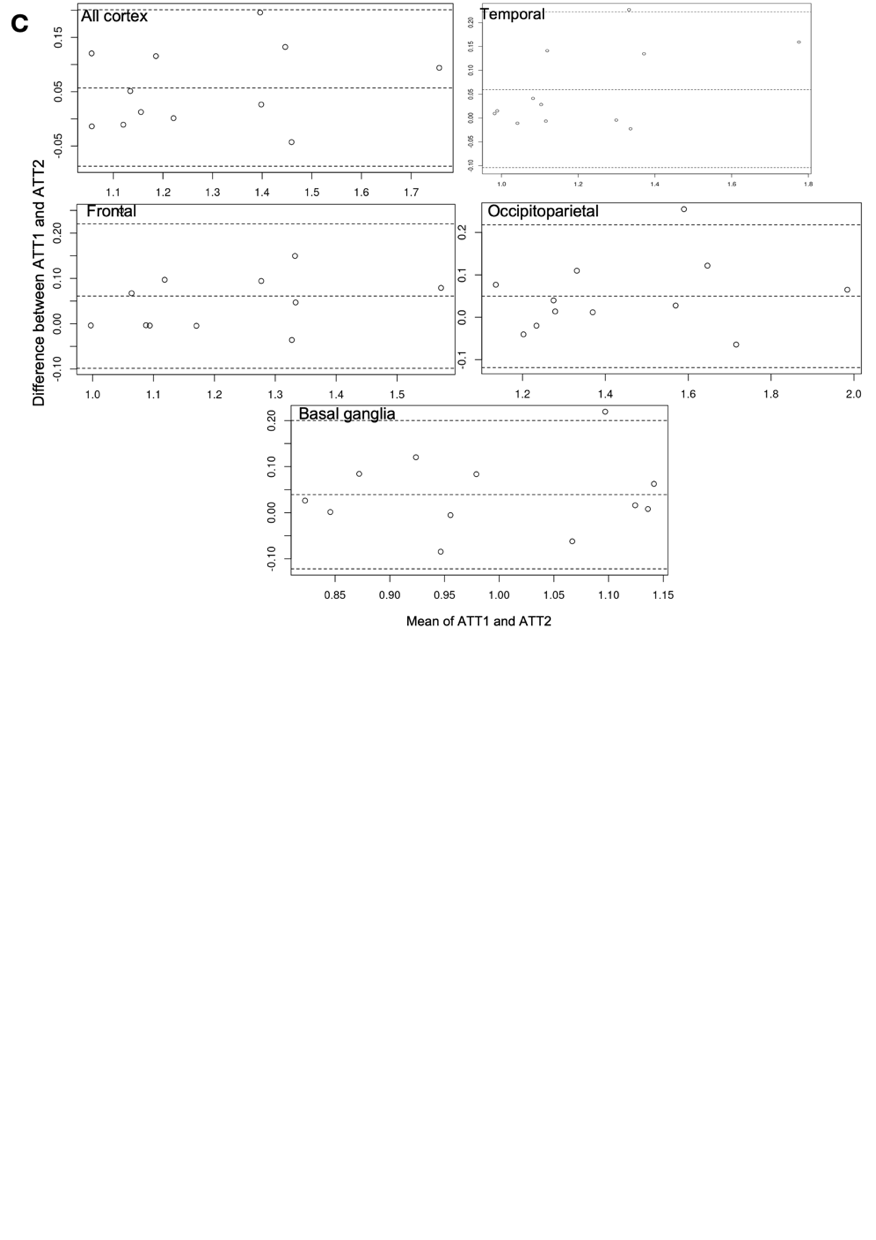

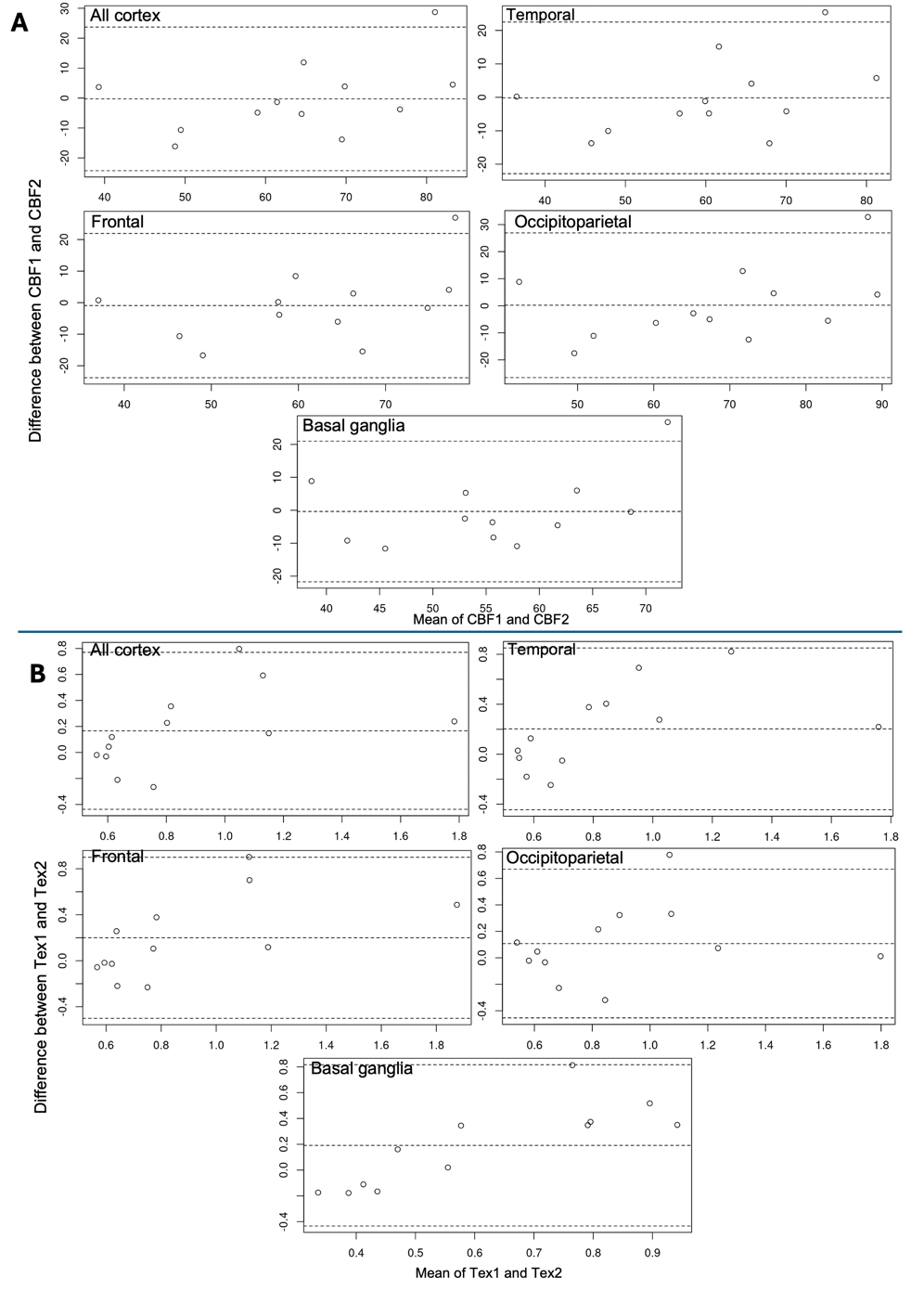
